# COVID-19 diagnosis prediction by symptoms of tested individuals: a machine learning approach

**DOI:** 10.1101/2020.05.07.20093948

**Authors:** Yazeed Zoabi, Noam Shomron

## Abstract

**Motivation:** Effective screening of SARS-CoV-2 enables quick and efficient diagnosis of COVID-19 and can mitigate the burden on healthcare systems. Prediction models that combine several features to estimate the risk of infection have been developed in hopes of assisting medical staff worldwide in triaging patients when allocating limited healthcare resources.

**Results:** We established a machine learning approach that trained on records from 51,831 tested individuals (of whom 4,769 were confirmed COVID-19 cases) while the test set contained data from the following week (47,401 tested individuals of whom 3,624 were confirmed COVID-19 cases). Our model predicts COVID-19 test results with high accuracy using only eight features: gender, whether age is above 60, known contact with an infected individual, and five initial clinical symptoms.

**Summary:** Overall, based on the nationwide data publicly reported by the Israeli Ministry of Health, we developed a model that detects COVID-19 cases by simple features accessed by asking basic questions. Our framework can be used, among other considerations, to prioritize testing for COVID-19 when allocating limited testing resources.

**Availability:** All data used in this study was retrieved from the Israeli Ministry of Health website.

## 1 Introduction

The novel coronavirus disease 2019 (COVID-19) pandemic caused by the newly emerged SARS-CoV-2 is a critical and urgent threat to global health. The outbreak in early December 2019 in the Hubei province of the People’s Republic of China has spread worldwide. As of May 2020, the overall number of patients confirmed to have the disease has exceeded 3,580,000 in more than 180 countries, the number of people infected is probably much higher, and more than 250,000 people have died from COVID-19. ^1^

This pandemic continues to challenge medical systems worldwide in many aspects, including sharp increases in demands for hospital beds and critical shortages in medical equipment, while many healthcare workers have themselves been infected. Thus, the capacity for immediate clinical decisions and effective usage of healthcare resources is crucial. The most validated diagnosis test for COVID-19, using reverse transcriptase polymerase chain reaction (RT-PCR), is currently in shortage in developing countries. This contributes to increased infection rates and delays critical preventive measures.

Effective screening enables quick and efficient diagnosis of COVID-19 and can mitigate the burden on healthcare systems. Prediction models that combine several features to estimate the risk of infection have been developed in hopes of assisting medical staff worldwide in triaging patients when allocating limited healthcare resources. These models use features such as computer tomography (CT) scans ^2-5^, information available at hospital admission including clinical symptoms ^6^, and laboratory tests.^7^

In Israel, all diagnostic laboratory tests for COVID-19 are performed according to criteria determined by the Israeli Ministry of Health. While subject to change, these currently include the presence and severity of clinical symptoms, possible exposure to confirmed patients, geographical area, the risk of complications if infected, and other factors.^8^

## 2 Methods

### Study Data and Features

The Israeli Ministry of Health recently publicly released data of individuals who were tested for SARS-CoV-2 via RT-PCR assay of a nasopharyngeal swab ^9^ The dataset contains initial records, on a daily basis, for all citizens tested for COVID-19 nationwide. In addition to the test date and result, various information is available, including clinical symptoms, gender and a binary indication as to whether the tested individual is above age 60 years. Based on this data, we developed a model that predicts COVID-19 test results using eight features: gender, whether age is above 60, known contact with an infected individual, and five initial clinical symptoms.

The training set consisted of records from 51,831 tested individuals (of whom 4,769 were confirmed COVID-19 cases), from the period March 22^th^, 2020 through March 31^st^, 2020. The test set contained data from the following week, April 1^nd^ through April 7^th^(47,401 tested individuals of whom 3,624 are confirmed COVID-19 cases).

The following list describes each of the features used by the model:

A. Basic information:

1. Gender (male/female).
2. Age ≥ 60 (true/false)
B. Symptoms:

3. Cough (true/false).
4. Fever (true/false).
5. Sore throat (true/false).
6. Shortness of breath (true/false).
7. Headache (true/false).
C. Other information:

8. Known contact with a confirmed COVID-19 individual (true/false).

**Table 1.**
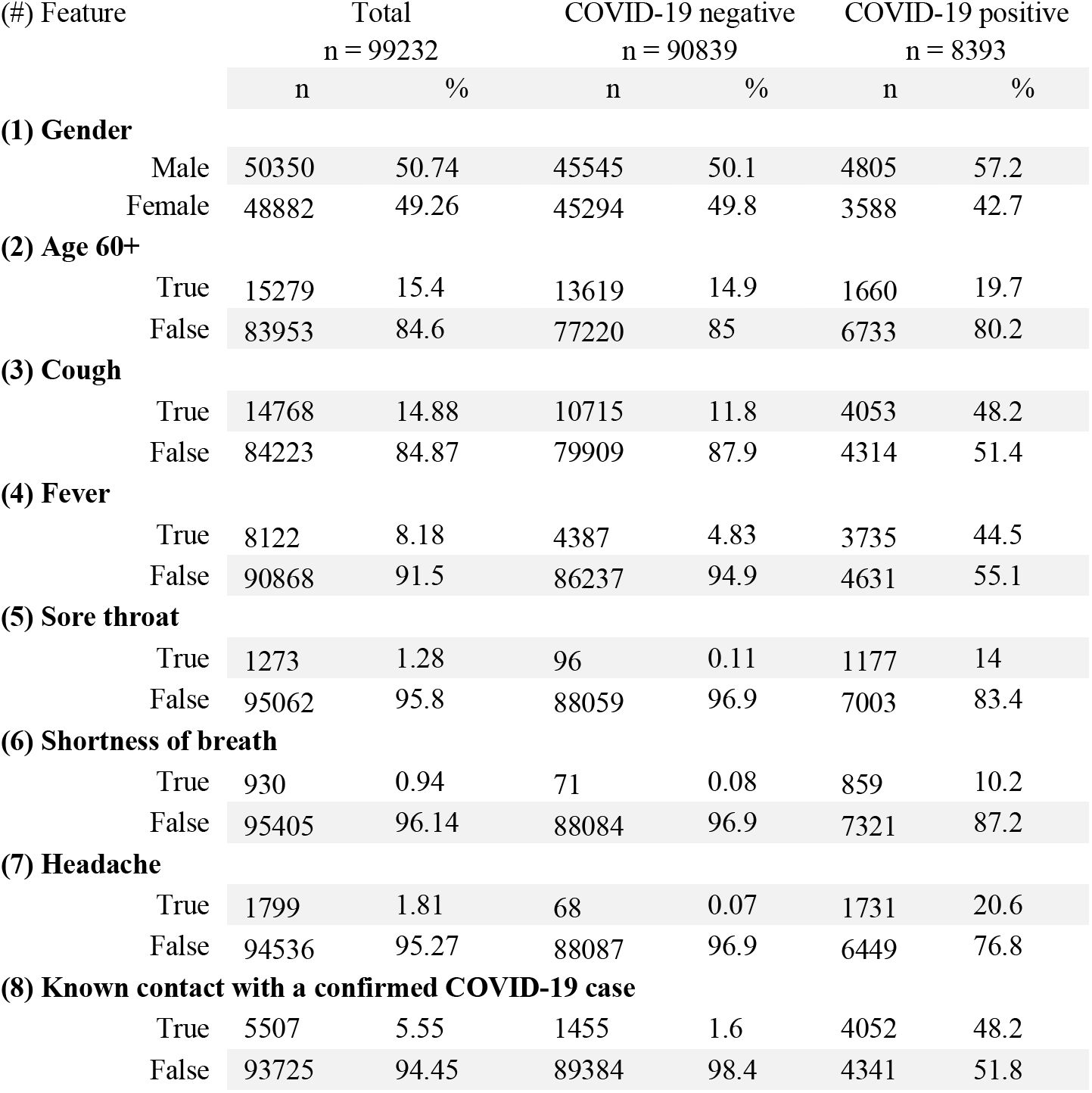
Characteristics of the dataset and the features used by the model in this study.

| (#) Feature | Total<br>n = 99232 |  | COVID-19 negative<br>n = 90839 |  | COVID-19 positive<br>n = 8393 |  |
| --- | --- | --- | --- | --- | --- | --- |
|  | n | % | n | % | n | % |
| **(1) Gender** |  |  |  |  |  |  |
| Male | 50350 | 50.74 | 45545 | 50.1 | 4805 | 57.2 |
| Female | 48882 | 49.26 | 45294 | 49.8 | 3588 | 42.7 |
| **(2) Age 60+** |  |  |  |  |  |  |
| True | 15279 | 15.4 | 13619 | 14.9 | 1660 | 19.7 |
| False | 83953 | 84.6 | 77220 | 85 | 6733 | 80.2 |
| **(3) Cough** |  |  |  |  |  |  |
| True | 14768 | 14.88 | 10715 | 11.8 | 4053 | 48.2 |
| False | 84223 | 84.87 | 79909 | 87.9 | 4314 | 51.4 |
| **(4) Fever** |  |  |  |  |  |  |
| True | 8122 | 8.18 | 4387 | 4.83 | 3735 | 44.5 |
| False | 90868 | 91.5 | 86237 | 94.9 | 4631 | 55.1 |
| **(5) Sore throat** |  |  |  |  |  |  |
| True | 1273 | 1.28 | 96 | 0.11 | 1177 | 14 |
| False | 95062 | 95.8 | 88059 | 96.9 | 7003 | 83.4 |
| **(6) Shortness of breath** |  |  |  |  |  |  |
| True | 930 | 0.94 | 71 | 0.08 | 859 | 10.2 |
| False | 95405 | 96.14 | 88084 | 96.9 | 7321 | 87.2 |
| **(7) Headache** |  |  |  |  |  |  |
| True | 1799 | 1.81 | 68 | 0.07 | 1731 | 20.6 |
| False | 94536 | 95.27 | 88087 | 96.9 | 6449 | 76.8 |
| **(8) Known contact with a confirmed COVID-19 case** |  |  |  |  |  |  |
| True | 5507 | 5.55 | 1455 | 1.6 | 4052 | 48.2 |
| False | 93725 | 94.45 | 89384 | 98.4 | 4341 | 51.8 |

### Statistical Analysis

Predictions were generated using a gradient-boosting machine model built with decision-tree base-learners ^10^. Gradient boosting is widely considered state of the art in predicting tabular data^11^ and is used by many successful algorithms in the field of machine learning ^12^. As suggested by previous studies ^13^, missing values were inherently handled by the gradient-boosting predictor^14^. We used the gradient-boosting predictor trained with the LightGBM ^15^ Python package.

To identify the principal features driving model prediction, SHAP (SHapley Additive eXplanations) values ^16^ were calculated. These values are suited for complex models such as artificial neural networks and gradient-boosting machines ^17^. Originating in game theory, SHAP values partition the prediction result of every sample into the contribution of each constituent feature value. This is done by estimating the difference between models with subsets of the feature space. By averaging across samples, SHAP values estimate the contribution of each feature to overall model predictions.

## 3 Results

For the prospective tests set, the model predicted with 0.90 auROC (area under the receiver operating curve) with 95% CI: 0.892-0.905 (Figure 1.a). Possible working points are: 87.3% sensitivity and 72% specificity, or 85.7% sensitivity and 79% specificity.

**Figure 1.**
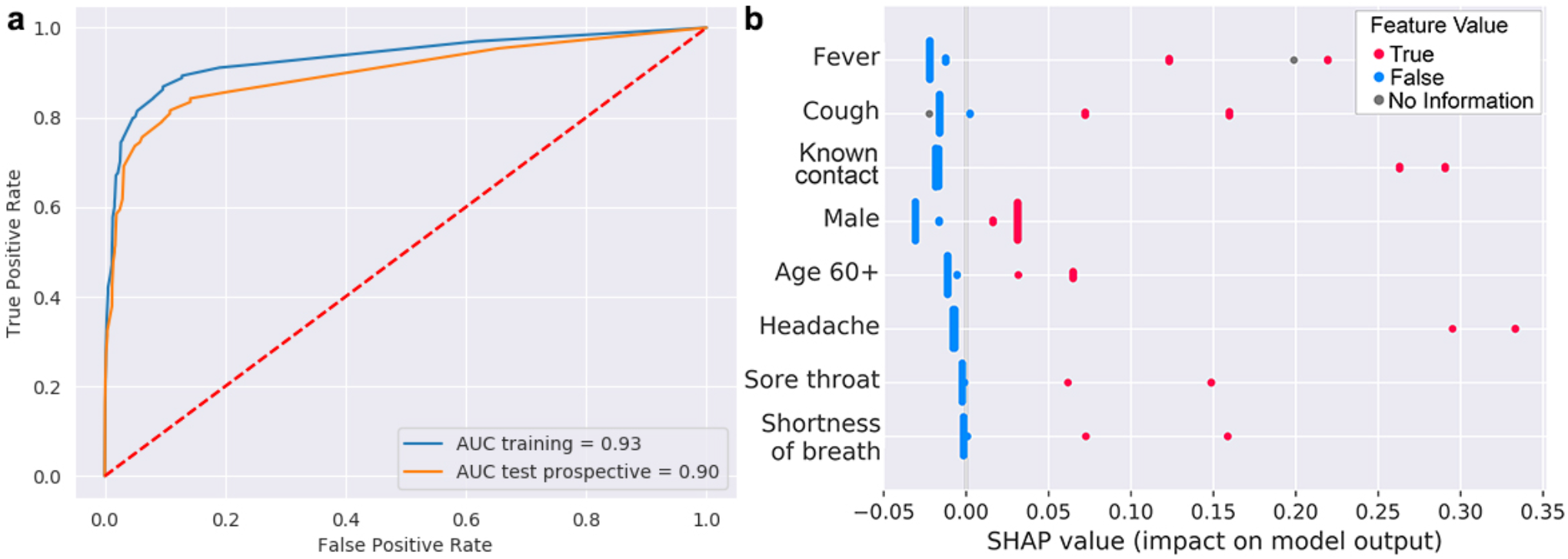
**a**. ROC curves of the predictive model. The blue line reflects training and testing via cross-validation. The orange line reflects testing the model on the prospective dataset. **b. SHapley Additive exPlanations** (SHAP) summary plots for COVID-19 diagnosis prediction show the SHAP values for the most important features of the model. Features in the summary plots (y-axis) are organized by their mean absolute SHAP values (x-axis), which represent the importance of that feature in driving the classifier’s prediction. Values of those features for each patient (i.e. fever) are colored by their relative value.

Our framework also provides ranking of the most important features that were used to define the decisions (Figure 1.b). Presenting with fever and cough were key features in predicting contraction of the disease. As expected, close contact with a confirmed COVID-19 individual was also an important feature, thus corroborating the disease’s high transmissibility ^18^. In addition, ‘male’ gender was revealed as a predictor of a positive result by the model, concurring with the observed gender bias ^19,20^

## 4 Discussion

This research is not without shortcomings. We relied on the data reported by the Israeli Ministry of Health, which has limitations and biases. For instance, symptom reporting was more comprehensive in the positive test result group and validated with a directed epidemiological effort ^21^. This can be reflected by the percentage of COVID-19 positive patients from the overall individuals positive for each symptom, with which we identified features with biased reporting (headache 96.2%, sore throat 92.3% and shortness of breath 92.4%) and symptoms with balanced reporting (cough 27.4% and fever 45.9%). We should also note that all symptoms were self-reported, and a negative value for a symptom can also mean that the symptom was not reported. If we train and test our model while filtering out symptoms of high bias in advance, we get an auROC of 0.862 with a slight change in the SHAP summary plot (Supplementary Figure 1).

However, we hope that readers will appreciate the rapid rate at which the pandemic scenario has evolved over the past weeks and understand the limitations of this research while also acknowledging that unusual times call for unusual solutions. We highlight the need for more robust data to complement our framework while also acknowledging the fact that self-reporting of symptoms is always subject to bias. As the COVID-19 pandemic progresses, it is crucial for public organizations and associations to continue recording and sharing robust data with the scientific community that is eager to contribute to the ongoing scientific effort.

Overall, based on the nationwide data reported by the Israeli Ministry of Health, we developed a model that detects COVID-19 cases by simple features accessed by asking eight basic questions. Our framework can be used, among other considerations, to prioritize testing for COVID-19 when allocating limited testing resources.

## Data Availability

All data used in this study was retrieved from the Israeli Ministry of Health website.

https://data.gov.il/dataset/covid-19

## Acknowledgements

We thank Professor David Gurwitz, Shomron lab members Artem Danilevsky and Guy Shapira for their comments on this work. Y.Z. is partially supported by the Edmond J. Safra Center for Bioinformatics at Tel-Aviv University.

## Supplementary Information

**Supplementary Figure 2.**
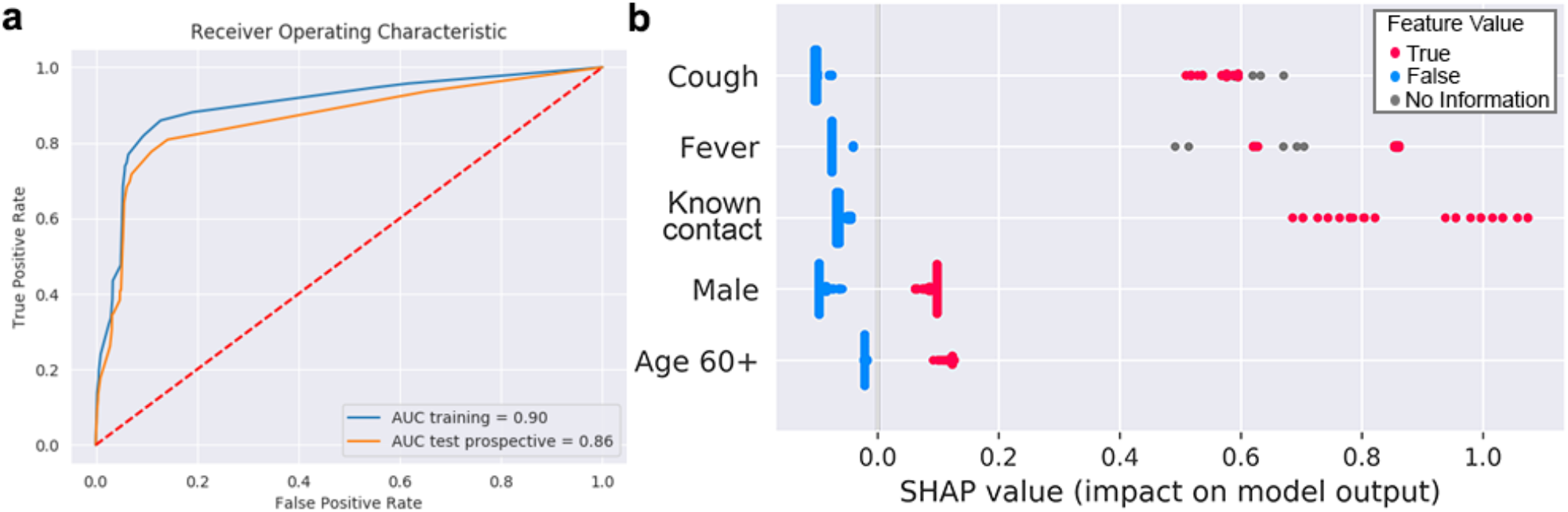
a. ROC curves and b. SHAP summary plots for training and testing using only balanced features.

